# A Phenome-Wide Association Study (PheWAS) of Genetic Risk for C-Reactive Protein in Children of European Ancestry: Results From the ABCD Study

**DOI:** 10.1101/2024.08.30.24312857

**Authors:** Sara A. Norton, Aaron J. Gorelik, Sarah E. Paul, Emma C. Johnson, David AA Baranger, Jayne L Siudzinski, Zhaolong Adrian Li, Erin Bondy, Hailey Modi, Nicole R. Karcher, Tamara Hershey, Alexander S. Hatoum, Arpana Agrawal, Ryan Bogdan

## Abstract

**BACKGROUND:** C-reactive protein (CRP) is a moderately heritable marker of systemic inflammation that is associated with adverse physical and mental health outcomes. Identifying factors associated with genetic liability to elevated CRP in childhood may inform our understanding of variability in CRP that could be targeted to prevent and/or delay the onset of related health outcomes.

**METHODS:** We conducted a phenome-wide association study (PheWAS) of genetic risk for elevated CRP (i.e. CRP polygenic risk score [PRS]) among children genetically similar to European ancestry reference populations (median analytic n = 5,509) from the Adolescent Brain and Cognitive Development^SM^ (ABCD) Study. Associations between CRP PRS and 2,377 psychosocial and neuroimaging phenotypes were estimated using independent mixed effects models. *Post hoc* analyses examined whether: (1) covarying for measured body mass index (BMI) or removing the shared genetic architecture between CRP and BMI altered phenotypic associations, (2) sex moderated CRP PRS associations, and (3) associations are unconfounded by assortative mating or passive gene-environment correlations (using a within-family analyses). Multiple testing was adjusted for using Bonferroni and false discovery rate (FDR) correction.

**RESULTS:** Nine phenotypes were positively associated with CRP PRS after multiple testing correction: five weight- and eating-related phenotypes (e.g. BMI, overeating), three phenotypes related to caregiver somatic problems (e.g. caregiver somatic complaints), as well as weekday video watching (all *p*s = 1.2 x 10^-7^ - 2.5 x 10^-4^, all *pFDR*s = 0.0002 - 0.05). No neuroimaging phenotypes were associated with CRP PRS (all *p*s = 0.0003 - 0.998; all *pFDR*s = 0.08 - 0.998) after correction for multiple testing. Eating and weight-related phenotypes remained associated with CRP PRS in within-family analyses. Covarying for BMI resulted in largely consistent results, and sex did not moderate any CRP PRS associations. Removing the shared genetic variance between CRP and BMI attenuated all relationships; associations with weekday video watching, caregiver somatic problems and caregiver report that the child is overweight remained significant while associations with waist circumference, weight, and caregiver report that child overeats did not.

**DISCUSSION:** Genetic liability to elevated CRP is associated with higher weight, eating, and weekday video watching during childhood as well as caregiver somatic problems. These associations were consistent with direct genetic effects (i.e., not solely due to confounding factors like passive gene-environment correlations) and were independent of measured BMI. The majority of associations with weight and eating phenotypes were attributable to shared genetic architecture between BMI and inflammation. The relationship between genetics and heightened inflammation in later life may be partially attributable to modifiable behaviors (e.g. weight and activity levels) that are expressed as early as childhood.

**Highlights:**

- Genetic risk for elevated CRP is associated with weight, eating, and screen time during childhood
- Associations were largely independent of measured BMI and were not moderated by sex
- Removing shared genetic variance between CRP and BMI attenuated all associations
- Modifiable childhood behaviors may mediate genetic liability to elevated inflammation

## 1. Introduction

As medical problems characterized by chronic systemic inflammation are the leading causes of global mortality, there have been extensive efforts to identify how individual differences in inflammation emerge (Furman et al., 2019). To date, the majority of this work has focused on C-reactive protein (CRP), an acute phase protein produced in the liver, which plays a crucial role in the nonspecific immune response by activating the complement system (Sproston & Ashworth, 2018). Following infection or injury, CRP levels elevate rapidly and typically decline upon resolution. However, sustained elevation of CRP may indicate chronic inflammation, which has been associated with a variety of adverse health outcomes. Indeed, meta-analyses, including of longitudinal data, have linked elevated CRP to a wide variety of physical and mental health outcomes (e.g., cardiovascular disease, obesity, cancer, type 2 diabetes, depression, schizophrenia, all-cause mortality; Ellulu et al., 2017; Emerging Risk Factors Collaboration, 2010; Ridker et al, 2003; Trichopoulos et al., 2006; Orsolini et al., 2022; Fond et al., 2018; Bernabe-Ortiz et al., 2022) and their respective risk factors (e.g., stress exposure, low socioeconomic status, low nutritional diets, physical inactivity; Johnson et al., 2013; Muscatell et al., 2020; Neale et al., 2016; Edwards & Loprinzi, 2018; Fedewa et al., 2017). Theories have speculated that individual differences in inflammation may initially emerge during early life as a result of genetic influences and early experiences that induce proinflammatory tendencies within cells, as well as behavioral proclivities (e.g., vigilance, impaired self-regulation, lifestyle choices associated with elevated BMI) that may directly and indirectly amplify inflammatory signaling throughout life (Miller et al., 2011, Nusslock et al., 2024). Understanding the phenotypic correlates of genetic liability to CRP during childhood may help identify biologic, behavioral, and environmental mechanisms that may contribute to heightened inflammation in later life and could be targeted through preventative efforts.

Building upon well-powered (e.g., n > 1,000) twin studies showing that CRP is moderately heritable (i.e., 0.43-0.53; Neijts et al., 2013; Rahman et al., 2009; Sas et al., 2017), a recent genome-wide association study (GWAS) of over 575,000 adults whose genetic ancestry resembles that of European reference populations identified 266 independent loci associated with CRP (Said et al., 2022). This GWAS characterized the genetic architecture of CRP and revealed evidence of genetic overlap with multiple inflammation-related health problems (e.g., depression, type 2 diabetes, ischemic heart disease, coronary artery disease, atherosclerosis, macular degeneration, and cancer), highlighting the role of genetically-associated differences in CRP to disease risk. The results generated from this GWAS provide a novel resource to investigate how genetic liability to elevated CRP in adulthood is correlated with other phenotypes across the lifespan, including during early life prior to the typical onset of associated health conditions. More specifically, the individual effect estimates of each variant from this study can be additively combined in the form of a polygenic risk score (PRS) to estimate an individual’s polygenic liability to CRP levels in an independent sample (Sugrue & Desikan, 2019).

Phenome-wide association studies (PheWAS) explore associations between a variable of interest (such as genetic risk, quantified via PRS) and hundreds or thousands of phenotypes spanning multiple domains. Effects that survive correction for multiple testing may replicate previously discovered genotype-phenotype associations or identify novel associations that may further drive the development of new hypotheses and theory (Bastarache et al., 2022; Bush et al., 2016; Wang et al., 2021). Here, we conducted a psychosocial and neuroimaging PheWAS of genetic risk for elevated CRP (i.e., CRP PRS) among children genetically similar to European reference populations (median analytic n = 5,509) who completed the baseline session of the Adolescent Brain and Cognitive Development^SM^ (ABCD) Study (Volkow et al., 2018). Three additional sets of analyses were conducted to examine the influence of body mass index (BMI), sex, and potential genetic confounds. *First*, given CRPs moderate-high phenotypic and genetic correlations with BMI (Choi et al., 2013; Ligthart et al., 2018) and the lack of inclusion of BMI in the discovery GWAS used to generate our CRP PRS, additional analyses examined whether covarying for measured BMI or removing shared genetic variance between CRP and BMI (i.e., generating CRP- minus-BMI PRS) altered phenotypic associations. *Second*, given sex-specific differences in circulating CRP (Cartier et al., 2009; Khera et al., 2005), we tested whether sex moderated CRP PRS phenotypic associations. *Third*, and finally, as genetic confounds (e.g., assortative mating, wherein individuals are more likely to select a partner who is genetically or phenotypically similar to themselves; Horwitz et al., 2023) can inflate GWAS test statistics and polygenic score associations (Okbay et al., 2022), we conducted a within-family analysis to assess whether any of the significant associations arising from either the CRP PRS or CRP-BMI PRS analyses are consistent with direct genetic effects. Ultimately, understanding how phenotypes in childhood are associated with genetic risk for CRP in later life may help elucidate the dynamic nature of health and disease across the lifespan. In particular, this approach may highlight potential modifiable mechanisms that could be leveraged during early life to prevent and/or delay the development of inflammation-related health conditions (Sas et al., 2017).

## 2. Materials and Methods

### 2.1. Participants

The ongoing longitudinal Adolescent Brain and Cognitive Development^SM^ (ABCD) Study recruited 11,879 children ages 9-11 (born 2005 - 2009) at baseline from 21 research sites across the United States to study health and development from middle childhood to early adulthood (Saragosa-Harris et al., 2022; Volkow et al., 2018). It includes a family-based component in which twin (n=2108), triplet (n=30), non-twin siblings (n=1,589), and singletons (n=8,148) were recruited. Baseline session data (collected 2016 – 2018) were drawn from data releases 3.0 (genetic), 4.0 (psychosocial), and 5.0 (neuroimaging) hosted at the National Institute of Mental Health Data Archive (NDA; https://nda.nih.gov/).

Participants who were not genetically similar to reference populations of European genomic ancestry (see Genetic Data section below) were excluded from our analyses due to the lack of well powered (1,000 participants or more in a community sample) ancestry-specific discovery GWAS of CRP in non-European ancestries and the relatively uninformative and low predictive utility of PRS when applied across ancestries (Martin et al., 2019). After further excluding individuals with missing covariate data, our final analytic sample consisted of n = 5,556 children of predominantly European genomic ancestry (assessed via principal components analysis) with baseline study data. Due to missing phenotypic data, the average analytic n per phenotype was 5,012 (median n = 5,509, range = 120 – 5,556, see **Supplemental Tables 1-9** for the n of each phenotype).

### 2.2. Phenotypes

Consistent with our prior PheWAS (Gorelik et al., 2023; Paul et al., in press), all released ABCD phenotypes were reviewed for inclusion according to: 1) relevance (e.g., we removed administrative items [e.g., measurement device], redundancy [e.g., excluding t-scored data and using raw data]), and 2) missingness and frequency variability (i.e., continuous phenotypes were required to have ≥ 100 participants with non-missing values; categorical variables required ≥ 100 endorsements/category). When applicable, missing data were re-coded to 0 (e.g., substance use questions that were not asked following a response that the child had not heard of the substance). Otherwise, all missing values were coded as missing (e.g., distress related to the presence of psychotic-like experiences was coded as missing in participants who reported no psychotic-like experiences). Data were also evaluated for skew, and variables with skew ≥ |1.96| had data points winsorized to +/- 3 SDs. Variables that still contained high skew after winsorization were subjected to a rank-based inverse normal transformation. All data were triple checked by multiple investigators for relevance, variability, and accurate re- coding.

Data were separated into psychosocial (n = 1,273 variables; **Supplemental Table 1**) and neural phenotypes (n = 1,104; **Supplemental Table 2**). Psychosocial data were grouped into the following 8 broad categories: 1) cognition (n = 14), 2) screen time (n = 18), 3) demographics (n = 27), 4) substance (n = 48), 5) culture/environment (n = 113), 6) physical health (n = 174), 7) family mental health (n = 239), and 8) child mental health (n = 640; **Supplemental Table 1**). Neural phenotypes included the following domains; further details can be found in (Casey et al., 2018; Hagler et al., 2019): 1) structural (n = 244), 2) resting state functional connectivity (RSFC; n = 339), 3) (diffusion tensor imaging (DTI): (white matter tract: fractional anisotropy [FA] and mean diffusivity [MD] n = 74), 4) restriction spectrum imaging (RSI): (restricted normalized isotropic diffusion [RNI] and restricted normalized directional anisotropic diffusion [RND]; n = 422), and global variables (e.g. whole brain characteristics such as total volume, surface area, etc.; n = 25). Neural phenotypes were derived using FreeSurfer segmentation (subcortical volumes; Dale et al., 1999), the Desikan-Killianry atlas (cortical thickness and surface area; Desikan et al., 2006), white matter AtlasTrack (FA, MD; Basser et al., 1994; Hagler et al., 2009), and Gordon networks (RSFC; Gordon et al., 2016). No task-related functional magnetic resonance imaging (fMRI) data were examined due to low test-retest reliability that precludes individual differences research (Elliott et al., 2020).

### 2.3. Genetic Data and CRP Genetic Risk Indices

Saliva samples were genotyped on the Smokescreen array (Baurley et al., 2016) by the Rutgers University Cell and DNA Repository (now SAMPLED; https://sampled.com/). Genotyped calls were aligned to GRC37 (hg19). The Rapid Imputation and COmputational PIpeLIne for Genome-Wide Association Studies (RICOPILI; Lam et al., 2020) was used to perform quality control (QC) on the 11,099 individuals with available ABCD Study phase 3.0 genotypic data, using RICOPILI’s default parameters. The 10,585 individuals who passed QC checks were subjected to principal component analysis (PCA) in RICOPILI to map the genomic ancestry of these individuals to the 1000 Genomes reference panel, resulting in a PCA-selected European-ancestry subset of 5,556 individuals. Only individuals of European ancestry were analyzed (due to the absence of available large-scale GWAS of CRP in non-European ancestry samples) as there is poor predictive utility across ancestries which may lead to erroneous conclusions (e.g., false negatives; Martin et al., 2019). The TOPMed imputation reference panel was used for imputation (Taliun et al., 2021). Imputation dosages were converted to best-guess hard-called genotypes, and only SNPs with Rsq > 0.8 and MAF > 0.01 were kept for PRS analyses.

Polygenic risk scores for CRP (CRP PRS) were generated for ABCD participants using PRS-CS (Ge et al., 2019), a Bayesian method that uses continuous shrinkage (CS) priors to weight SNP effect sizes. We used effect sizes from the largest (n = 575,531 European descent) currently available GWAS of CRP (Said et al., 2022). We used the ‘auto’ function of PRS-CS, allowing the software to learn the global shrinkage parameter from the data, and the number of MCMC (Markov chain Monte Carlo) iterations was set at 10,000 and the number of burn-ins was set at 5,000. After deriving SNP weights using PRS-CS, we used PLINK 1.9’s (Chang et al., 2015) -- score command to produce PRS in the ABCD sample.

### 2.4. Statistical Analysis

#### Primary Analysis

Numeric data (including the PRS) were z-scored prior to analysis. Associations between CRP PRS and phenotypes were estimated using independent linear mixed effects models in the lme4 R software package (Bates et al., 2015); the lmer() function was used for continuous outcomes, and the generalized glmer() function was used for dichotomous outcomes. All non-imaging models were nested by site and family ID while imaging models were nested by scanner and family ID to account for the non-independence of these data.

#### Covariates

Fixed effect covariates for all analyses included: the first 10 ancestral principle components, age, and sex. Sex was removed for models where the outcome was a sex-specific phenotype (e.g., “Have you noticed a deepening of your voice?” was only asked of boys). Imaging models also included MRI manufacturer and global brain metrics (for region-specific analyses) as fixed effect covariates; for DTI, RSI, and RSFC models, mean head motion was also included.

#### Multiple Testing Correction

Bonferroni and False Discovery Rate (FDR) corrections were applied across all psychosocial phenotypes, as well as neuroimaging phenotypes by domain, consistent with our prior work (Baranger et al., 2024). Bonferroni correction was used to minimize false positives and identify the most robust associations that are most likely to replicate and generalize across samples, while FDR correction was used to minimize false negatives that may be meaningful but undetected with the more stringent Bonferroni correction. Bonferroni alpha levels for each domain were: (0.05/1,273) = 3.928 × 10^-5^ for psychosocial phenotypes; (0.05/244) = 2.049 × 10^-4^ for neural structural phenotypes; (0.05/339) = 1.433 × 10^-4^ for RSFC phenotypes; (0.05/74) = 6.757 × 10^-4^ for DTI phenotypes; (0.05/211) = 2.370× 10^-4^ for both RNI and RND phenotypes; and (0.05/25) = 2.000 x 10^-3^ for global brain phenotypes.

#### Post-hoc Analyses

***1) BMI.*** First, given phenotypic and genetic correlations between CRP and BMI (Ligthart et al., 2018), the CRP PRS PheWAS was repeated covarying additionally for BMI. We examined whether significant associations (other than BMI) seen in the primary analyses remained associated with CRP PRS; we then also conducted a full PheWAS of all other phenotypes to assess whether the BMI-adjusted models revealed any new associations. Second, we removed genetic variance that is shared between CRP and BMI. To this end, we applied multi-trait conditional and joint analysis (mtCOJO; Zhu et al., 2018) to condition the CRP GWAS summary statistics (Said et al., 2022) on the summary statistics of the largest BMI GWAS (Yengo et al., 2018). This approach generated GWAS-based summary statistics for CRP that are unique from BMI, from which we generated a CRP-BMI PRS where the genetic variance shared with BMI was removed. Significant CRP PRS phenotypic associations from primary analyses were re-tested with CRP-BMI PRS; we also re-ran full PheWAS across all variables to test whether any novel associations emerged (and corrected for multiple testing as above).
***2) Sex.*** Given sex differences in CRP and related health outcomes (Khera et al., 2009; Nari et al., 2020), we tested whether the associations between CRP PRS and phenotypes were moderated by sex. To this end, we regressed each phenotype on the interaction between CRP PRS and sex while accounting for main effects of sex and CRP PRS. We also included interactions between covariates and sex (e.g., ancestry PC1 x sex) and covariates and PRS, consistent with recommendations to adequately account for potential confounds in interaction analyses (Keller, 2014).
***3) Within-Family Analysis.*** For any significant CRP PRS associations with phenotypes that may have within-family variability (e.g., BMI, but not caregiver self-reports, which would not vary across siblings), we conducted follow-up within-family analyses to assess whether our identified associations may plausibly represent direct genetic effects (Selzam et al., 2019). If effects remain significant within-family (as opposed to just between-family), this would suggest that these associations are not confounded by population stratification, assortative mating, passive gene-environment correlations, or other potential population-level confounds (though it should be noted that active and evocative genetic effects could still influence within-family variation in PRS effects; (Brumpton et al., 2020; Howe et al., 2022). For these analyses, we included both the family mean PRS and each sibling’s deviation from their family mean PRS as predictors in a mixed-effect model, as done previously (Gorelik et al., 2023; Selzam et al., 2019).

## 3. Results

### 3.1. Primary Analyses

#### 3.1.1. Psychosocial Phenotypes

Three weight-related phenotypes (measured BMI, caregiver report that child is overweight, and measured waist circumference) were significantly positively associated with CRP PRS after Bonferonni correction (all |β|s 0.0629 – 0.0707; all *p*s = 1.2 x 10^-7^ – 1.9 x 10^-6^; **Figure 1; Supplemental Table 3**). An additional three child phenotypes (measured weight, caregiver report that child overeats, weekday video watching) and three caregiver phenotypes (eye-problems not corrected by glasses, caregiver self-report of somatic problems on the ASR-scale and DSM-5 scales) were positively associated with CRP PRS following FDR correction (all |β|s = 0.045 – 0.050 ; all *p*s = 8.6 x 10^-5^ - 2.5 x 10^-4^ , pFDRs = 0.01 - 0.05). One phenotype, caregiver report of drug use (excluding alcohol or nicotine; ASR question 6), showed a trending association after FDR adjustment (β = 0.036; *p* = 5.9 x 10^-4^, *p*FDR = 0.07;).

**Figure 1.**
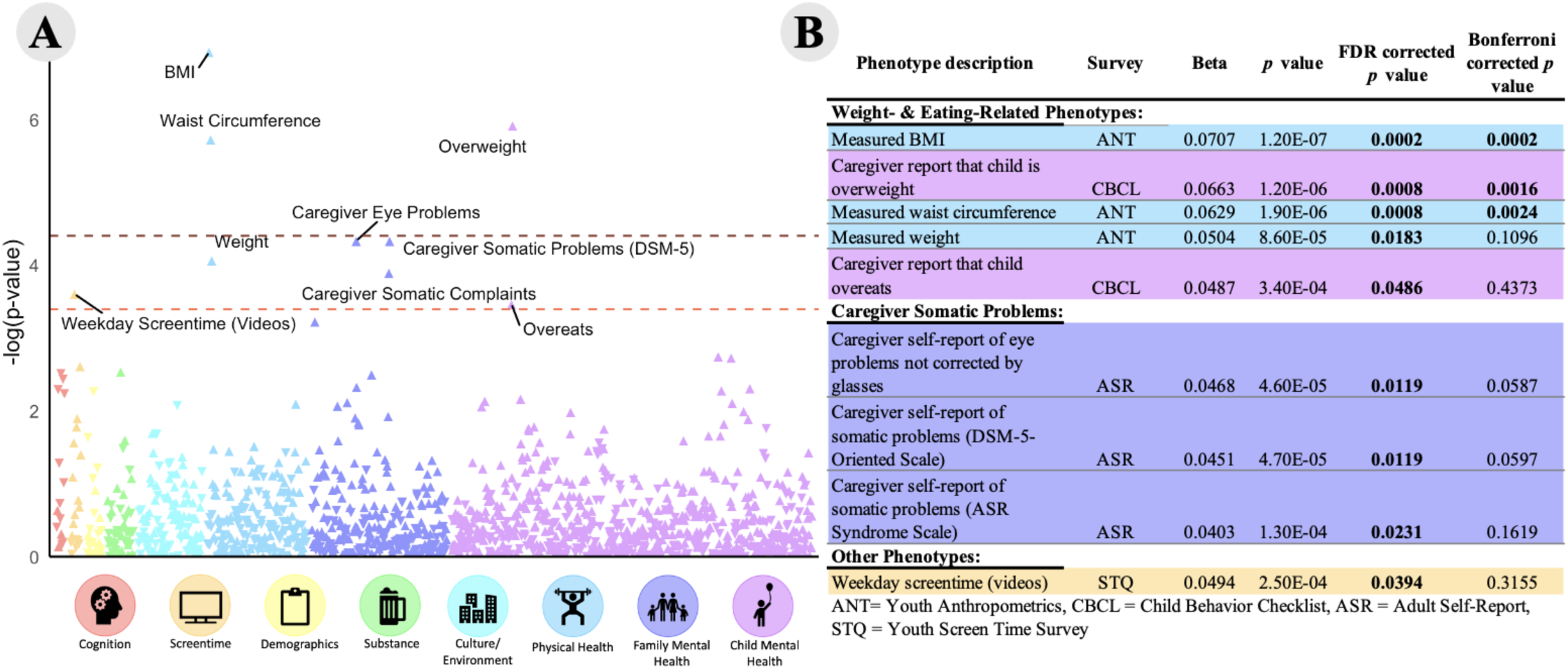
CRP PRS was associated with 5 weight and eating-related phenotypes, 3 caregiver somatic problems, and 1 screentime phenotype. **(A) A** Manhattan plot of all 1,273 psychosocial phenotypes. Phenotypes above the dark red dotted line are significant after Bonferroni correction; phenotypes above the light red dotted line are significant after FDR correction. (B) Table of significant associations with p values, uncorrected p values, and corrected p values. Corrected p values < .05 are bolded, p and p values for all phenotypes can be found in **Supplemental Table 3**.

#### 3.1.2. Neural Phenotypes

No neural phenotypes were associated with CRP PRS after Bonferroni or FDR adjustment within modalities (all |β|s = 7.77 x 10^-5^ – 0.047; all *p*s = 3.9 x 10^-4^ – 0.998 pFDR = 0.08 – 0.998; ;Supplemental Tables 4-9).

### 3.2. Post-hoc Testing

#### 3.2.1. Covarying for Measured BMI

Covarying for measured BMI resulted in largely consistent results. CRP PRS remained significantly associated with all 8 psychosocial phenotypes (excluding BMI) identified in primary analyses (all |β|s = 0.040 – 0.067; all ps = 9.1 x 10^-7^ – 3.5 x 10^-4^, pFDRs = 7.28 x 10^-6^ – 0.0004 ; **Supplemental Table 10**). No other phenotypes became significantly associated with CRP PRS after multiple testing correction.

#### 3.2.2. mtCOJO CRP-BMI PRS

The three caregiver somatic complaints (i.e., caregiver self-report of eye problems not corrected by glasses and the two caregiver self-reports of somatic problems) identified in primary analyses were also significantly associated with CRP-BMI PRS (all |β|s = 0.03 – 0.04; all *p*s= 0.0008 – 0.003, *p*FDRs = 0.005 – 0.006; **Supplemental Table 11**). Caregiver report that the child is overweight and weekday video watching were significant after FDR correction, but not Bonferroni correction (|β|s = 0.03; *p* = 0.01, pFDR = 0.02). Three phenotypes related to anthropometrics and eating (i,e., waist circumference, weight, and parent report that the child overeats) that were significant in the main analysis were not significantly associated with the conditioned CRP-BMI PRS (all *p*s > 0.2). Associations with CRP-BMI PRS were attenuated for all psychosocial phenotypes (e.g., caregiver eye problems for CRP PRS was β = 0.047, SE = 0.012; for CRP-BMI β = 0.038, SE = 0.011). One neural phenotype, fiber segmentation of the right parahippocampal cingulum, was significant after FDR correction (β = -0.035, *p* = 5.18 x 10^-5^, *p*FDR = 0.011; **Supplemental Table 12**); in our primary CRP PRS analyses, this association was trending after multiple testing correction (*p*FDR = 0.09; **Supplemental Table 7**). No other neural phenotypes became significantly associated with CRP-BMI PRS after multiple testing corrections (**Supplemental Tables 13-17**).

**3.2.22. CRP PRS x Sex Interaction.** Sex did not significantly moderate any CRP PRS-phenotype association (all |β|s = 3.15 x 10^-16^ – 3.05; all *p*s = 0.0007 – 0.999, *p*FDR = 0.05 – 0.999; **Supplemental Tables 18-24**)

**3.2.3. Within-family Analysis.** *Post-hoc* within-family analyses were conducted for all Bonferroni or FDR-corrected significant associations but excluding variables that would have the same value for all children in a family (e.g., caregiver self-reports). All of the psychosocial phenotypes, with the exception of the screentime phenotype (β = 0.02, p = 0.30), remained significant (all |β| = 0.035 – 0.042, all *p*s = 0.01 – 0.02 ; **Figure 2; Supplemental Table 25**).

**Figure 2.**
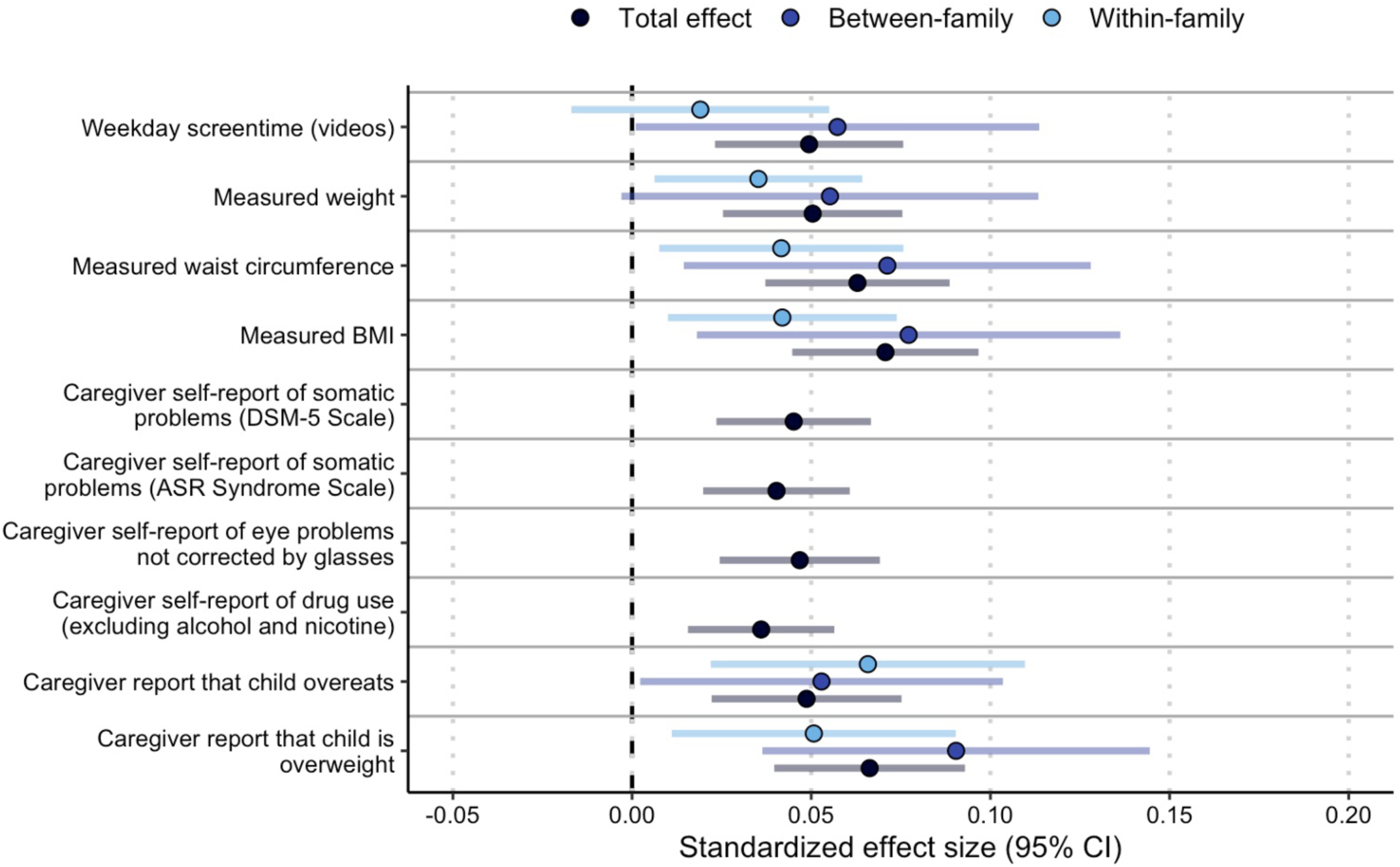
β coefficients and 95% confidence intervals for the nine phenotypes that survived FDR correction for multiple testing and one phenotype that was trending toward significance in the CRP PRS PheWAS of psychosocial phenotypes. Total effect is the effect in the full sample; between- and within-family effects were run in a sample restricted to sibling groups. Caregiver self-report phenotypes were not included in the within/between family analysis, as answers are expected to be the same for most sibling groups.

## 4. Discussion

Our PheWAS of CRP PRS in middle/late childhood revealed that genetic liability to elevated CRP in adulthood is associated with child weight and eating-related phenotypes (BMI, waist circumference, weight, and caregiver report that the child is overweight and overeats), screen time (child weekday video watching), and caregiver somatic complaints (somatic problems, eye problems not corrected by glasses). That most of these associations remained significant in our post hoc within-family analyses suggests these may arise from direct genetic effects or evocative gene-environment correlations (e.g., children’s genetically influenced behavior evokes a parental response to encourage their children to eat more) as opposed to assortative mating, ancestral stratification, or passive gene-environment correlations (**Figure 2**). Associations were independent of measured BMI, but the majority of associations with weight and eating phenotypes were attributable to the shared genetic associations between CRP and BMI. Taken together, these data suggest that genetic liability to elevated inflammation in adulthood manifests as heavier weight, overeating, and weekday video watching during middle/late childhood. Indeed, it may be that preventative and policy efforts aimed at weight management and activity during childhood may potentially reduce inflammation-related health problems in later life, particularly in those with heightened genetic liability to inflammation. As the participants of the ABCD Study continue to age, this will continue to be an invaluable sample in which to characterize protective factors against inflammation-related health outcomes.

## Weight and Eating

CRP PRS was most strongly associated with higher child weight- and eating-related phenotypes (**Figure 1**). Circulating CRP has been unequivocally linked to heightened BMI (e.g., being overweight or obese) through meta-analyses, and elevated CRP is observed in both overweight and obese adults (Visser et al., 1999) and adolescents (Konstantinos et al., 2013). Consistent with evidence that adipose tissue overexpresses pro-inflammatory cytokines in overweight and obese individuals (Ellulu et al, 2017), heightened weight and adiposity are hypothesized to be causal factors contributing to chronic elevations in systemic inflammation. Supportive of this causal interpretation, weight loss following lifestyle or surgical intervention is associated with a dose-dependent decrease in CRP (Selvin et al., 2007).

Our study raises the possibility that higher weight and related behaviors (i.e., overeating and sedentary behavior) during childhood are potential mechanisms through which genetic liability to heightened inflammation in adulthood emerges. These findings hint at potential gene-behavior correlations undergirding inflammation in childhood wherein genetic liability to inflammation in later life influences weight-related phenotypes that are expressed during middle childhood that could be directly tested with longitudinal data across the lifespan and non-human animal experiments. Consistent with this interpretation, when partialling out the shared genetic associations of CRP and BMI (via mtCOJO analyses), the unique polygenic risk for CRP was no longer associated with the majority of weight-related phenotypes.^1^ However, the caregiver report of child overeating remained associated with CRP PRS, even after partialling out the genetic associations shared with BMI. Interestingly, unlike the appetite-suppressing effects of acute infection, elevated basal levels of inflammation have been observed in adolescents who experience loss of control (LOC) eating (i.e., eating where there are feelings of being unable to stop eating), even when accounting for adiposity (Shank et al., 2017). While some evidence suggests that inflammation may induce broad elevations in impulsivity (Gassen et al., 2019), we found no evidence that polygenic liability to CRP was associated with broader impulsivity-related phenotypes (e.g. none of the UPPS-P impulsive behavior scale items included in the psychosocial analysis were significant). Collectively, our data suggest that genetic propensity to heightened CRP in adulthood may act through weight-related phenotypes in childhood.

## Screen Time

Genetic liability to elevated CRP was associated with greater time spent watching videos on weekdays. Notably, this association was robust to the inclusion of BMI as a covariate, and independent of genetic liability to BMI. Although not significant after multiple testing correction, there were nominal associations (uncorrected p < .05) between CRP PRS and other screen time measures (n = 5 out of 18). These screen time measures may reflect increased sedentary behavior. It is well known that in adults regular physical activity has anti-inflammatory^1^The discovery GWAS of CRP that was used to generate CRP PRS did not account for BMI due to concerns about inducing collider bias as well as prior evidence that the heritability of CRP is not modified when accounting for measured BMI and that prior genome-wide significant loci associated with CRP are not genome-wide significant in BMI GWAS (Said et al., 2022; Sas et al., 2017).

properties and physical activity and cardiorespiratory fitness are inversely associated with CRP levels (Albert et al., 2004; Church et al., 2002; Geffken et al., 2001; LaMonte et al., 2002; Pitsavos et al., 2003). Indeed, adults with higher activity levels have lower CRP than less active individuals (Plaisance & Grandjean, 2006), and randomly controlled trials of exercise suggest that adherence to an exercise program reduces CRP, with reductions in BMI accounting for only a small portion of this exercise-induced change in CRP (Fedewa et al., 2017). However, the relationship between exercise and CRP is less clear in children. As none of our 3 phenotypes related to physical activity were even nominally significant (**Supplemental Table 3)**, the association between sedentary behavior, physical activity, and genetic liability to CRP in children warrants further investigation. If genetic liability to elevated CRP in adulthood is associated with child sedentary behavior, this could represent a mechanism with high preventative impact for reducing inflammation-related disease later in life.

## Caregiver Somatic Complaints

Three phenotypes describing caregiver self-reported somatic complaints (eye-problems not corrected by glasses, caregiver self-report of somatic problems on the ASR-scale and DSM-5 scales) were significantly associated with CRP PRS across primary and post-hoc analyses. As caregivers in the ABCD study are likely to be biological parents of the participants (4,792 of 5,556 [86%] caregivers in this sample identified as the child’s mother) and therefore genetically related to the child, children and caregivers are very likely to have similar CRP PRS (i.e., children with high CRP PRS are more likely to have a caregiver with a high CRP PRS), these somatic complaints may reflect lifetime effects of high, genetically-driven levels of CRP. As the ABCD cohort continues to age, it will be of interest to see how these somatic complaints may emerge and manifest in individuals with high CRP PRS through late childhood and adolescence.

## Neuroimaging

No neuroimaging phenotypes were associated with polygenic liability to inflammation in our primary analyses. However, one restricted spectrum imaging phenotype, restricted normalized isotropic diffusion (RNI) in the right parahippocampal cingulum fiber tract, was negatively associated with CRP-BMI PRS. RNI has been theorized to track neuroinflammation, reflected by increased isotropic intracellular water presence in proliferating glial cells (White et al., 2013). Based on this, the direction of effect we observed suggests that elevated polygenic liability to CRP that is independent of BMI (CRP-BMI PRS) is associated with less putative neuroinflammation in the parahippocampal cingulum tract. As central and peripheral levels of inflammation are presumed to be correlated (Felger et al., 2020), and CRP-BMI PRS estimates genetic liability to elevated peripheral inflammation (i.e. CRP), this direction of effect is opposite of what would be expected. Another possible interpretation of RNI data is that it, at least partially, tracks normal neurodevelopment: higher RNI has been associated with greater age in the ABCD Study (Palmer et al., 2022), which might reflect increased dendritic sprouting and/or neuronal density. Therefore, our findings, which controlled for age, might reflect delayed white matter maturation associated with a BMI-independent CRP PRS. In support of this idea is the observation that children with chronic inflammatory conditions, such as inflammatory bowel disease, often have delayed pubertal onset (Amaro & Chiarelli, 2020; Ballinger et al., 2003; Grob & Zacharin, 2020), although none of the assessed genetic risk indices of CRP were associated with phenotypes assessing pubertal development (e.g., pubertal developmental scale) in our analyses.

## Limitations

The results of this study should be interpreted in the context of its limitations. *First,* this study was limited to children whose genetic ancestry resembles that of European reference populations due to the lack of CRP GWASs in other ancestries and evidence that applying GWAS results to other ancestries can result in erroneous associations that may further drive ancestral health disparities (Martin et al., 2019). As such, it is unknown whether these results may generalize to other ancestries. Given extensive efforts to broaden the ancestral diversity of genetic association research (Fatumo et al., 2022), we are hopeful that this question can be addressed in the future as adequately powered GWAS in other ancestral populations emerge. *Second,* while the large sample size and breadth of phenotypic assessment in the ABCD Study permitted a PheWAS approach, this approach also induces a significant multiple testing burden that may have resulted in false negative associations. We attempted to address this by including FDR multiple testing correction, which provides a more lenient threshold than Bonferroni multiple testing correction, as well as reporting all associations in **Supplemental Tables**. However, it remains possible that non-significant results may reflect false negatives. *Third,* CRP PRS were based on a GWAS of circulating CRP levels in adults (Said et al., 2022). It is possible that the genetic architecture of CRP varies across development, which may attenuate associations during childhood (Ferrucci & Fabbri, 2018; Li et al., 2023; Stumper et al., 2020). *Fourth,* we found no evidence that sex moderates associations between polygenic liability to CRP and our examined phenotypes; as sex differences in CRP between men and women do not emerge until after puberty (Shanahan et al., 2013), it is possible that our lack of CRP PRS x sex interactions is attributable to the young age of our sample and that such associations may be revealed as children progress into adolescence, as reproductive hormones are known to impact immune functioning (Shanahan et al., 2013; Stumper et al., 2020). *Finally,* our PheWAS was constrained by phenotypes measured in the ABCD study, which unfortunately does not include any measure of circulating inflammation levels (e.g. serum CRP or IL-6, WBC).

## Conclusion

Polygenic liability for elevated CRP in later life is associated with higher weight-related and screentime phenotypes among children as well as increased somatic complaints among their caregivers. These findings highlight potentially modifiable factors, such as weight and activity-related behaviors, through which genetic liability to elevated CRP may manifest. Preventative efforts and policies encouraging healthy eating and activity (e.g. providing healthy food and time for physical activity) among children may help attenuate susceptibility to inflammation and age-related disease conferred by genetic liability to CRP.

## Data Availability

All data produced in the present study are available upon reasonable request to the authors.

## Acknowledgements

We are thankful to families who have participated in the ABCD Study as well as study staff and investigators.

